# Computer vision-based assessment of motor functioning in schizophrenia: Use of smartphones for remote measurement of schizophrenia symptomatology

**DOI:** 10.1101/2020.07.20.20158287

**Authors:** Anzar Abbas, Vijay Yadav, Emma Smith, Elizabeth Ramjas, Sarah B. Rutter, Caridad Benavides, Vidya Koesmahargyo, Li Zhang, Lei Guan, Paul Rosenfield, Mercedes Perez-Rodriguez, Isaac R. Galatzer-Levy

## Abstract

**Introduction:** Motor abnormalities have been shown to be a distinct component of schizophrenia symptomatology. However, objective and scalable methods for assessment of motor functioning in schizophrenia are lacking. Advancements in machine learning-based digital tools have allowed for automated and remote ‘digital phenotyping’ of disease symptomatology. Here, we assess the performance of a computer vision-based assessment of motor functioning as a characteristic of schizophrenia using video data collected remotely through smartphones.

**Methods:** 18 patients with schizophrenia and 9 healthy controls were asked to remotely participate in smartphone-based assessments daily for 14 days. Video recorded from the smartphone front-facing camera during these assessments was used to quantify head movement through a pre-trained computer vision model. The ability of head movement measurements to distinguish between patients and healthy controls as well as their relationship to schizophrenia symptom severity as measured through traditional clinical scores was assessed.

**Results:** A logistic regression demonstrated that head movement was a significant predictor of schizophrenia diagnosis (*p* < 0.05). Linear regression between head movement and clinical scores of schizophrenia symptom severity showed that head movement has a negative relationship with schizophrenia symptom severity (*p* < 0.05), primarily with negative symptoms of schizophrenia.

**Conclusions:** Remote, smartphone-based assessments were able to capture meaningful visual behavior for computer vision-based objective measurement of head movement. The measurements of head movement acquired were able to accurately classify schizophrenia diagnosis and quantify symptom severity in patients with schizophrenia.

## 1 Introduction

Motor abnormalities have been shown to be a characteristic trait of schizophrenia, are present even in antipsychotic-naive patients, predate the onset of psychosis, and have been demonstrated in at-risk populations and in unaffected relatives.^1–6^ Indeed, psychomotor retardation underlies core negative symptomatology of schizophrenia such as the demonstration of blunted facial affect and emotional withdrawal.^7–11^ Though there have been many attempts to diagnose schizophrenia and quantify symptom severity using neurobiology and behavior,^12–14^ little work has been done to objectively measure motor dysfunction to characterize the disorder in clinical research,^15–17^ which could be of particular relevance in assessment of negative symptoms.

Recent advances in the mechanistic understanding of negative symptomatology have led to a number of promising pharmacological and cognitive treatments for negative symptoms of schizophrenia.^18–21^ Such initiatives are important given the lack of FDA-approved treatments for negative symptoms.^22^ However, reliable and change-sensitive measures of negative symptomatology to assess the efficacy of these treatments are sparse ^23–26^. With motor dysfunction being an underlying factor of negative symptomatology, accurate measurement of motor functioning can allow for assessment of treatment efficacy during the evaluation of novel investigational treatments.

Several groups have successfully and accurately measured motor functioning as a characteristic of schizophrenia, though only in laboratory-settings ^27–31^. While laboratory-based methodologies increase sensitivity and reduce subjectivity in measurement of motor functioning compared to traditional assessments, they are limited in their utility as measurement tools in clinical research given the burden imposed on both patients and clinicians.

The use of ‘digital phenotyping’ or measurement of disease symptomatology using digital tools has shown significant promise towards objective, scalable, and remote measurement of central nervous system functioning.^32,33^ In schizophrenia, observable behavior associated with the disorder has been successfully quantified using advancements in machine learning. Examples of this include using digital signal and natural language processing methods to measure changes in verbal prosody and speech characteristics such as volume, fundamental frequency, pause characteristics, and sentiment^34–39^ and computer vision to quantify changes in facial expressivity and presence of blunted facial affect, a core negative symptom of schizophrenia.^34–38^ The use of visual data such as videos of patient behavior presents a promising avenue for digital phenotyping given the ubiquitous availability of smartphone cameras alongside advances in computer vision methodologies that can automate the processing and analysis of visual information.

In the current investigation, we examined the ability of head movement measured using computer vision from videos recorded during remote smartphone-based assessments to differentiate individuals with schizophrenia from healthy controls and determine disease severity by comparison of head movement with the Positive and Negative Syndrome Scale (PANSS), a commonly used clinical assessment tool for measurement of schizophrenia severity and the current ‘gold standard’ for assessment of antipsychotic treatment efficacy.^39^

## 2 Methods

Code for all methods and analysis presented in this manuscript is publicly available on GitHub: https://github.com/anzarabbas/ms_headmov_scz.

### 2.1 Participants

Patients meeting DSM-5 criteria for schizophrenia at Mount Sinai Health System Outpatient Psychiatry Clinics (*n* = 18; age µ = 48.1, σ = 13.1; 11 females) and healthy individuals from the community (*n* = 9; age µ = 39.8, σ = 10.4; 5 females) were consented to participate in a two-week observational study under approval of the Mount Sinai Program for the Protection of Human Subjects and its Institutional Review Board. Participants in the schizophrenia group were on a stable regimen of psychotropic medications, had no recent clinically meaningful change in schizophrenia symptomatology, and were clinically stable in that they were not expected to show significant changes in symptomatology over the course of the two-week observational study.

### 2.2 Data collection

#### 2.2.1 Positive and negative syndrome scale (PANSS)

The Positive and Negative Syndrome Scale(PANSS) was administered in-person by the study team on the first day of the study on both patients with schizophrenia and healthy control subjects. Healthy individuals were assessed to confirm an absence of schizophrenia symptomatology. Ten individuals from the schizophrenia group were also assessed on the last day of the study i.e. day 14. For those ten individuals, the average of the two scores were used for all subsequent analyses. Given the patients were clinically stable, the average of the two PANSS scores simply allowed for reduction in noise. From the PANSS, subscale scores were recorded for the positive symptom subscale (P-total), the negative symptom subscale (N-total), and general symptom subscale (G-total) in addition to all individual items within those subscales.

#### 2.2.2 Remote smartphone-based video assessments

All participants were asked to download the AiCure app (www.aicure.com) on either their personal smartphone or a smartphone provisioned to them by the study team. They were then trained by the study team on how to use the app to participate in remote assessments. The remote assessments triggered the participants to perform a brief 1-minute task where they were asked open-ended questions to which they responded freely while a video of their response was captured using the front-facing camera on the smartphone. The open-ended questions were neutral in nature and simply meant to elicit a free verbal response (e.g. “What have you been doing for the past few hours?” and “What are your plans for the rest of the day?”). The assessments were scheduled everyday for the 14 days of the study. The videos collected during these assessments were used to quantify head movement during the participants’ responses.

### 2.3 Measurements of head movement

The software library OpenFace^40–42^ was used to measure frame-wise head movement from the videos collected through the remote smartphone-based assessments. For each frame, the head’s position relative to the camera was calculated using pre-trained convolutional neural network-based computer vision models. The software provides a confidence score for every frame of video denoting the likelihood that it is accurately detecting a face; only frames with a confidence score of 80% or higher were used for downstream analyses. Framewise measurements of change in euclidean head position in the *x, y*, and *z* planes were calculated in millimeters as shown in the equation below. From the framewise measurements, mean head movement was calculated across all videos collected from a participant over the course of the study. All measurements of head movement were normalized between 0 and 1 before subsequent data analysis.

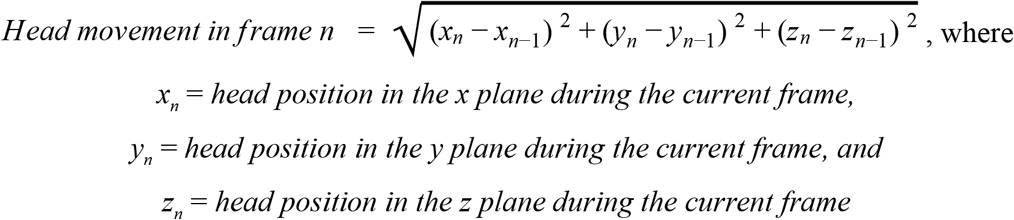

### 2.4 Data analysis

Using head movement data collected from all participants in both groups, a multiple logistic regression with maximum likelihood estimation was conducted to model the probability of an individual having a schizophrenia diagnosis based on measurements of head movement, with age and gender as additional predictors in the regression. Using data only from individuals with schizophrenia, separate linear regressions were conducted to determine the relationship between head movement and the PANSS subscale scores in addition to individual items in each of those subscales, with age and gender as confounding variables. The linear regressions were an exploratory analysis to determine the relationship between head movement and individual items on the PANSS and hence multiple comparisons correction was not conducted.

### 2.5 Data Availability

The raw video analyzed in this manuscript is considered Protected Health Information. It was not consented for public use and cannot be published. However, the first level of non-identifiable raw abstractions from that data as well as the derivations used to conduct all analyses will be provided alongside the code.

### 2.6 Code Availability

Code for all methods and analysis presented in this manuscript is publicly available on GitHub (https://github.com/anzarabbas/ms_headmov_scz).

## 3 Results

### 3.1 Logistic regression

Data from 9 healthy controls and 18 individuals with schizophrenia was used for the logistic regressions. Head movement, age, and gender were able to significantly differentiate healthy controls and individuals with schizophrenia (*LL* = −12.13; *Pseudo r*^*2*^ = 0.29; *p <* .05) with head movement as the only significant predictor (*O*.*R*. = 0.04 (95% CI: 0.00, 0.48); *z* = −2.27; *p* < .05) and age demonstrating a marginal effect (*O*.*R*. = 1.07 (95% CI: 0.99, 1.18); *z* = 1.65; *p* < .1) (Table 1).

**Table 1:** Results from multiple logistic regression of head movement, age, and gender with schizophrenia diagnoses.

| Variable | Coefficient | Standard Error | z | p-value |
| --- | --- | --- | --- | --- |
| Constant | -0.441 | 2.069 | -0.213 | 0.938 |
| Head motion | -5.421 | 2.392 | -2.267 | 0.023 |
| Age | 0.075 | 0.046 | 1.65 | 0.1 |
| Gender | -0.465 | 1.03 | -0.453 | 0.651 |

### 3.2 Linear regression

In the data collected from individuals with schizophrenia (*n* = 18), separate linear regressions were conducted with head movement, age, and gender as predictors of each individual item on the PANSS. The results from all of these regressions are detailed in Supplementary Tables 1, 2, and 3. A subset of the results from those regressions are outlined in Tables 2 and 3.

**Table 2:**
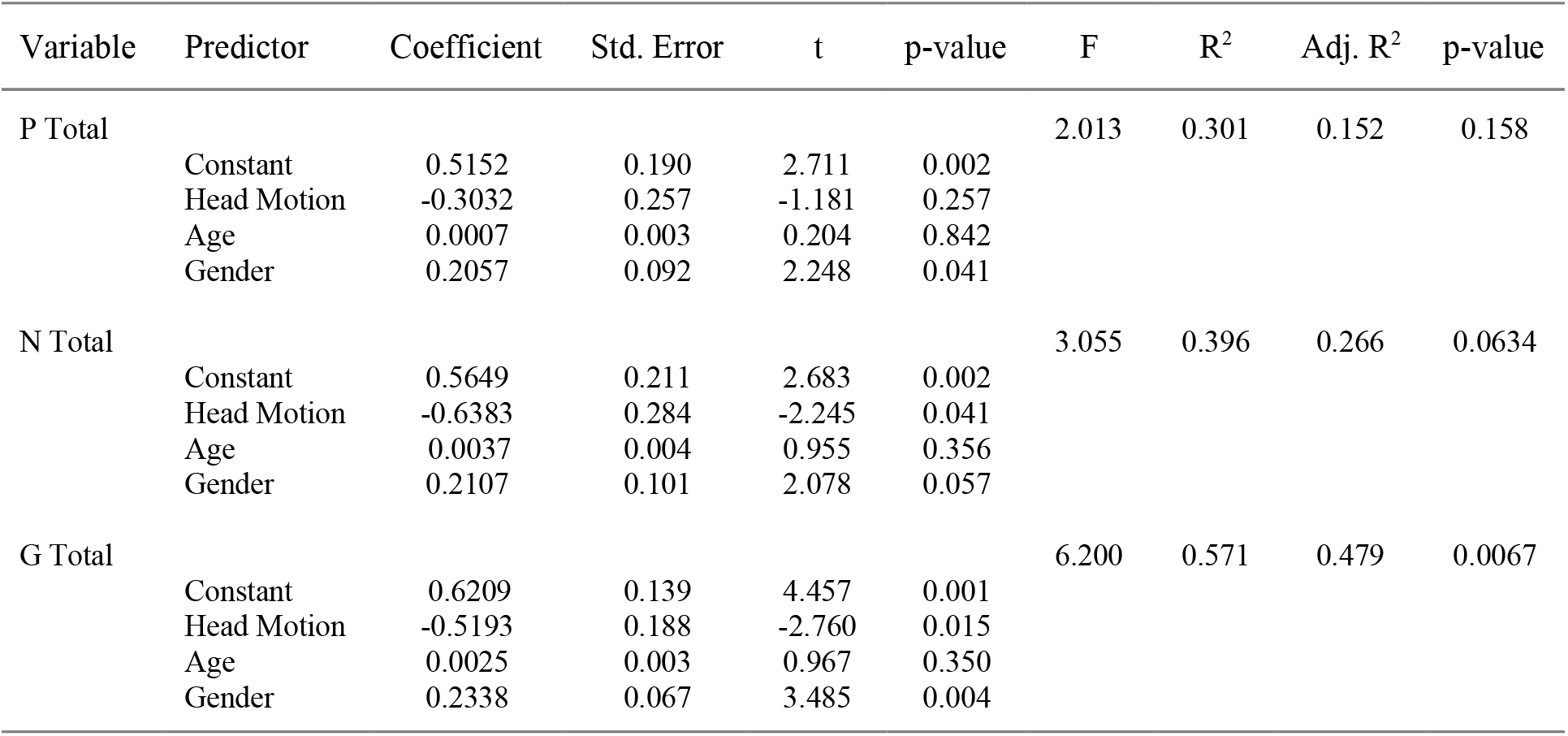
Results for the linear regressions conducted with head movement, age, and gender as predictors of PANSS subscale scores.

**Table 3:** A subset of the results for the linear regressions conducted with head movement, age, and gender as predictors of individual PANSS items.

| Dependent variable | Predictor | Coefficient | Std. Err. | t | p-value | F | R<sup>2</sup> | Adj. R<sup>2</sup> | p-value |
| --- | --- | --- | --- | --- | --- | --- | --- | --- | --- |
| **N1**<br>Blunted<br>Affect | Constant | 0.5700 | 0.226 | 2.517 | 0.025 | 3.064 | 0.396 | 0.267 | 0.0629 |
|  | Head Motion | -0.7412 | 0.306 | -2.424 | 0.029 |  |  |  |  |
|  | Age | 0.0041 | 0.004 | 0.997 | 0.336 |  |  |  |  |
|  | Gender | 0.2000 | 0.109 | 0.997 | 0.088 |  |  |  |  |
| **N2**<br>Emotional<br>Withdrawal | Constant | 0.5566 | 0.191 | 2.911 | 0.001 | 4.313 | 0.480 | 0.369 | 0.0237 |
|  | Head Motion | -0.7483 | 0.258 | -2.898 | 0.012 |  |  |  |  |
|  | Age | 0.0059 | 0.004 | 1.690 | 0.113 |  |  |  |  |
|  | Gender | 0.1604 | 0.092 | 1.742 | 0.103 |  |  |  |  |
| **N3**<br>Poor Rapport | Constant | 0.6747 | 0.237 | 2.843 | 0.013 | 2.395 | 0.339 | 0.198 | 0.112 |
|  | Head Motion | -0.7986 | 0.321 | -2.491 | 0.026 |  |  |  |  |
|  | Age | -0.0010 | 0.004 | -0.234 | 0.818 |  |  |  |  |
|  | Gender | 0.1294 | 0.114 | 1.133 | 0.276 |  |  |  |  |
| **G7**<br>Motor<br>Retardation | Constant | 0.6199 | 0.243 | 2.554 | 0.023 | 4.023 | 0.463 | 0.348 | 0.0294 |
|  | Head Motion | -1.0616 | 0.328 | -3.239 | 0.006 |  |  |  |  |
|  | Age | 0.0050 | 0.004 | 1.127 | 0.279 |  |  |  |  |
|  | Gender | 0.1160 | 0.117 | 0.992 | 0.338 |  |  |  |  |
| **G8**<br>Uncooperativeness | Constant | 0.7052 | 0.224 | 3.154 | 0.007 | 2.190 | 0.319 | 0.174 | 0.135 |
|  | Head Motion | -0.6850 | 0.302 | -2.269 | 0.040 |  |  |  |  |
|  | Age | -0.0019 | 0.004 | -0.455 | 0.656 |  |  |  |  |
|  | Gender | 0.1322 | 0.108 | 1.228 | 0.240 |  |  |  |  |
| **G12**<br>Lack of Judgement<br>and Insight | Constant | 0.4446 | 0.230 | 1.937 | 0.073 | 3.069 | 0.397 | 0.267 | 0.0626 |
|  | Head Motion | -0.7751 | 0.310 | -2.500 | 0.025 |  |  |  |  |
|  | Age | 0.0060 | 0.004 | 1.414 | 0.179 |  |  |  |  |
|  | Gender | -0.0687 | 0.111 | -0.622 | 0.544 |  |  |  |  |

When correcting for age and gender, head movement had a significant negative association with the N-total and G-total scores (*p* = 0.041 and *p* = 0.015 respectively) but not the P-total score (*p* = 0.257) (Table 2). Further, when correcting for age and gender, head movement was negatively associated with several individual items on the PANSS (Table 3): N1 blunted affect (*p* = 0.029), N2 emotional withdrawal (*p* = 0.012), N3 poor rapport (*p* = 0.026), G7 motor retardation (*p* = 0.006), G8 uncooperativeness (*p* = 0.040), and G12 lack of judgement and insight (*p* = 0.025).

## 4 Discussion

In the current investigation, we sought to test the hypothesis that motor functioning as measured by the amount of head movement during free behavior, quantified through convolutional neural network-based computer vision models, would serve as a digital marker of schizophrenia symptomatology. We further assessed the ability of digital tools, i.e. remote smartphone-based assessments that capture video of individuals’ behavior, to measure schizophrenia symptomatology in an accurate, scalable, and objective manner.

Findings demonstrate that head movement during free responses to scripted prompts delivered over a smartphone and recorded through the front-facing camera significantly differentiated healthy controls from individuals with schizophrenia (Table 1). Further, the exploratory results suggest that slowed head movement is a marker of negative symptomatology of schizophrenia including blunted affect, motor retardation, poor rapport, and emotional withdrawal (Table 2). Additionally, we found preliminary evidence that head movement is associated with uncooperativeness and lack of insight. While the interpretation of this relationship is less straightforward, this may reflect underlying cognitive deficits in schizophrenia associated with negative symptomatology. Our findings align with previously reported motor slowing in individuals with schizophrenia^43–46^ and are consistent with the notion that different types of motor symptoms may correlate with different aspects of psychotic symptoms.^47^

Furthermore, the results demonstrated that motor functioning can be objectively measured using data captured remotely, in the absence of a clinician or a clinical environment, and without the need for any specialized equipment or hardware beyond a smartphone. Video captured of an individual’s behavior during remote smartphone-based assessments was sufficient for measurement of head movement as a behavioral symptom of schizophrenia, and may be sufficient for computationally-based measurement of other behavioral symptoms such as changes in facial expressivity, vocal acoustics, and characteristics of speech. With tools emerging that are able to measure all such behavioral symptomatology using computer vision, digital signal processing, and machine learning ^34–38,48,48–53^, these behaviors may be measurable in the same manner that movement behavior was assessed in this investigation.

Abnormalities in motor functioning underlie observable symptomatology in several neurological and neuropsychiatric disorders including Major Depressive Disorder, ^54,55^ Parkinson’s Disease,^56^ and Autism Spectrum Disorder, ^57–59^ among others. Given the transdiagnostic nature of motor symptomatology, remote and scalable methods for its measurement may have utility for assessment of treatment response and/or motor side-effects of treatment in both clinical care and clinical research beyond just schizophrenia. With advancements in computationally-based measurement of other observable behavioral symptomatology such as facial expressivity, speech, and physiology, digital measurements could bring immense utility in the monitoring and diagnosis of mental and physical health using remote tools.

It is important to note key limitations of this study, in particular the multiple comparisons conducted on the modest sample size. With the primary objective of this study being to test the hypothesis that head movement is negatively correlated with schizophrenia diagnosis, the logistic regression provided compelling evidence towards rejection of the null hypothesis. The subsequent linear regressions conducted between head movement and all PANSS items served as a secondary and exploratory aim, allowing for determination of a relationship between head movement and specific schizophrenia symptomatology. With results from the analysis indicating relationships as would be hypothesized based on prior reports, we present them as preliminary findings with the ultimate goal of expanding such an experiment to a wider patient population, allowing for a more fine-grained analysis of head movement as a digital biomarker of schizophrenia symptomatology.

Finally, this investigation utilized open-source Python-based software, available to all researchers. As an additional measure, the code that implemented the open-source software for this investigation and subsequent analyses of results have been provided by the authors in the Methods section. This allows for the expansion of the experiment to a wider patient population as mentioned above and the independent validation of the computer vision-based measurement of head movement itself and its implementation in this investigation by other researchers in academic and clinical research, following an open-science framework for the development of digital tools for objective, accurate, and scalable measurement of disease symptomatology in both mental and physical health.

## 5 Conclusions

In this investigation, we demonstrate that head movement measured using computer vision from video captured remotely via smartphones demonstrates validity as a marker of schizophrenia and is a promising metric for negative symptom severity. Use of such technology in clinical care and clinical research settings could allow for accurate measurement of disease symptomatology and treatment response in a scalable and accessible manner, which can support development of novel treatments for schizophrenia and other mental and physical disorders that involve motor symptomatology.

## Acknowledgments

The authors appreciate the involvement of the clinical, research, and operations staff at both Mount Sinai and AiCure for the development, deployment, and implementation of the technology presented here and the participants who volunteered to be involved in the research.

## Supplementary Materials

**Supplementary Table 1:** Results for the multiple linear regressions conducted with head movement, age, and gender as predictors of individual items on the PANSS P subscale.

| Dependent variable | Predictor | Coefficient | Std. Err. | t | p-value | F | R<sup>2</sup> | Adj. R<sup>2</sup> | p-value |
| --- | --- | --- | --- | --- | --- | --- | --- | --- | --- |
| **P1**<br>Delusions | Constant | 0.2849 | 0.212 | 1.341 | 0.201 | 4.494 | 0.491 | 0.381 | 0.0208 |
|  | Head Motion | -0.4222 | 0.287 | -1.47 | 0.163 |  |  |  |  |
|  | Age | 0.0046 | 0.004 | 1 | 0.254 |  |  |  |  |
|  | Gender | 0.3453 | 0.102 | 1.190 | 0.005 |  |  |  |  |
|  |  |  |  | 3.376 |  |  |  |  |  |
| **P2**<br>Conceptual disorganization | Constant | 0.5376 | 0.236 | 2.274 | 0.039 | 3.928 | 0.457 | 0.341 | 0.0316 |
|  | Head Motion | -0.6582 | 0.319 | -2.06 | 0.058 |  |  |  |  |
|  | Age | 0.0014 | 0.004 | 2 | 0.750 |  |  |  |  |
|  | Gender | 0.3314 | 0.114 | 0.325 | 0.011 |  |  |  |  |
|  |  |  |  | 2.911 |  |  |  |  |  |
| **P3**<br>Hallucinatory behavior | Constant | 0.2612 | 0.385 | 0.678 | 0.509 | 0.469<br>6 | 0.091 | -0.103 | 0.708 |
|  | Head Motion | -0.0547 | 0.520 | -0.10 | 0.918 |  |  |  |  |
|  | Age | 0.0053 | 0.007 | 5 | 0.463 |  |  |  |  |
|  | Gender | 0.1843 | 0.185 | 0.755 | 0.337 |  |  |  |  |
|  |  |  |  | 0.994 |  |  |  |  |  |
| **P4**<br>Excitement | Constant | 0.4433 | 0.219 | 2.022 | 0.063 | 1.196 | 0.204 | 0.033 | 0.347 |
|  | Head Motion | 0.3482 | 0.296 | 1.176 | 0.259 |  |  |  |  |
|  | Age | -0.0039 | 0.004 | -0.96 | 0.353 |  |  |  |  |
|  | Gender | 0.0973 | 0.106 | 2 | 0.372 |  |  |  |  |
|  |  |  |  | 0.921 |  |  |  |  |  |
| **P5**<br>Grandiosity | Constant | 0.7971 | 0.271 | 2.946 | 0.011 | 0.796<br>5 | 0.146 | -0.037 | 0.516 |
|  | Head Motion | -0.4925 | 0.365 | -1.34 | 0.199 |  |  |  |  |
|  | Age | -0.0037 | 0.005 | 8 | 0.467 |  |  |  |  |
|  | Gender | 0.0041 | 0.130 | -0.74 | 0.975 |  |  |  |  |
|  |  |  |  | 8<br>0.032 |  |  |  |  |  |
| **P6**<br>Suspiciousness | Constant | 0.7946 | 0.214 | 3.721 | 0.002 | 2.847 | 0.379 | 0.246 | 0.0755 |
|  | Head Motion | -0.2764 | 0.288 | -0.95 | 0.354 |  |  |  |  |
|  | Age | -0.0026 | 0.004 | 8 | 0.517 |  |  |  |  |
|  | Gender | 0.2735 | 0.103 | -0.66 | 0.019 |  |  |  |  |
|  |  |  |  | 4<br>2.660 |  |  |  |  |  |
| **P7**<br>Hostility | Constant | 0.4090 | 0.295 | 1.387 | 0.187 | 0.654<br>6 | 0.123 | -0.065 | 0.593 |
|  | Head Motion | -0.5236 | 0.398 | -1.31 | 0.210 |  |  |  |  |
|  | Age | 0.0027 | 0.005 | 4 | 0.632 |  |  |  |  |
|  | Gender | 0.0339 | 0.142 | 0.489 | 0.815 |  |  |  |  |
|  |  |  |  | 0.239 |  |  |  |  |  |

**Supplementary Table 2:** Results for the multiple linear regressions conducted with head movement, age, and gender as predictors of individual items on the PANSS N subscale

| Dependent variable | Predictor | Coefficient | Std. Err. | t | p-value | F | R<sup>2</sup> | Adj. R<sup>2</sup> | p-value |
| --- | --- | --- | --- | --- | --- | --- | --- | --- | --- |
| **N1**<br>Blunted Affect | Constant | 0.5700 | 0.226 | 2.517 | 0.025 | 3.064 | 0.396 | 0.267 | 0.0629 |
|  | Head Motion | -0.7412 | 0.306 | -2.424 | 0.029 |  |  |  |  |
|  | Age | 0.0041 | 0.004 | 0.997 | 0.336 |  |  |  |  |
|  | Gender | 0.2000 | 0.109 | 0.997 | 0.088 |  |  |  |  |
| **N2**<br>Emotional Withdrawal | Constant | 0.5566 | 0.191 | 2.911 | 0.001 | 4.313 | 0.480 | 0.369 | 0.0237 |
|  | Head Motion | -0.7483 | 0.258 | -2.898 | 0.012 |  |  |  |  |
|  | Age | 0.0059 | 0.004 | 1.690 | 0.113 |  |  |  |  |
|  | Gender | 0.1604 | 0.092 | 1.742 | 0.103 |  |  |  |  |
| **N3**<br>Poor Rapport | Constant | 0.6747 | 0.237 | 2.843 | 0.013 | 2.395 | 0.339 | 0.198 | 0.112 |
|  | Head Motion | -0.7986 | 0.321 | -2.491 | 0.026 |  |  |  |  |
|  | Age | -0.0010 | 0.004 | -0.234 | 0.818 |  |  |  |  |
|  | Gender | 0.1294 | 0.114 | 1.133 | 0.276 |  |  |  |  |
| **N4**<br>Passive/apathetic social withdrawal | Constant | 0.3798 | 0.229 | 1.655 | 0.120 | 1.761 | 0.274 | 0.118 | 0.201 |
|  | Head Motion | -0.4349 | 0.310 | -1.403 | 0.182 |  |  |  |  |
|  | Age | 0.0052 | 0.004 | 1.236 | 0.237 |  |  |  |  |
|  | Gender | 0.1754 | 0.111 | 1.588 | 0.135 |  |  |  |  |
| **N5**<br>Difficulty in abstract thinking | Constant | 0.5441 | 0.332 | 1.638 | 0.124 | 1.546 | 0.249 | 0.088 | 0.247 |
|  | Head Motion | -0.0918 | 0.449 | -0.205 | 0.841 |  |  |  |  |
|  | Age | -0.0014 | 0.006 | 0.227 | 0.824 |  |  |  |  |
|  | Gender | 0.3444 | 0.160 | 2.153 | 0.049 |  |  |  |  |
| **N6**<br>Lack of spontaneity & flow of conversation | Constant | 0.5248 | 0.295 | 1.780 | 0.097 | 1.181 | 0.202 | 0.031 | 0.353 |
|  | Head Motion | -0.6833 | 0.398 | -1.716 | 0.108 |  |  |  |  |
|  | Age | 0.0018 | 0.005 | 0.330 | 0.746 |  |  |  |  |
|  | Gender | 0.1249 | 0.142 | 0.880 | 0.394 |  |  |  |  |
| **N7**<br>Stereotyped thinking | Constant | 0.4336 | 0.233 | 1.859 | 0.084 | 2.991 | 0.391 | 0.260 | 0.0668 |
|  | Head Motion | -0.5788 | 0.315 | -1.837 | 0.088 |  |  |  |  |
|  | Age | 0.0053 | 0.004 | 1.238 | 0.236 |  |  |  |  |
|  | Gender | 0.2580 | 0.112 | 2.297 | 0.038 |  |  |  |  |

**Supplementary Table 3:**
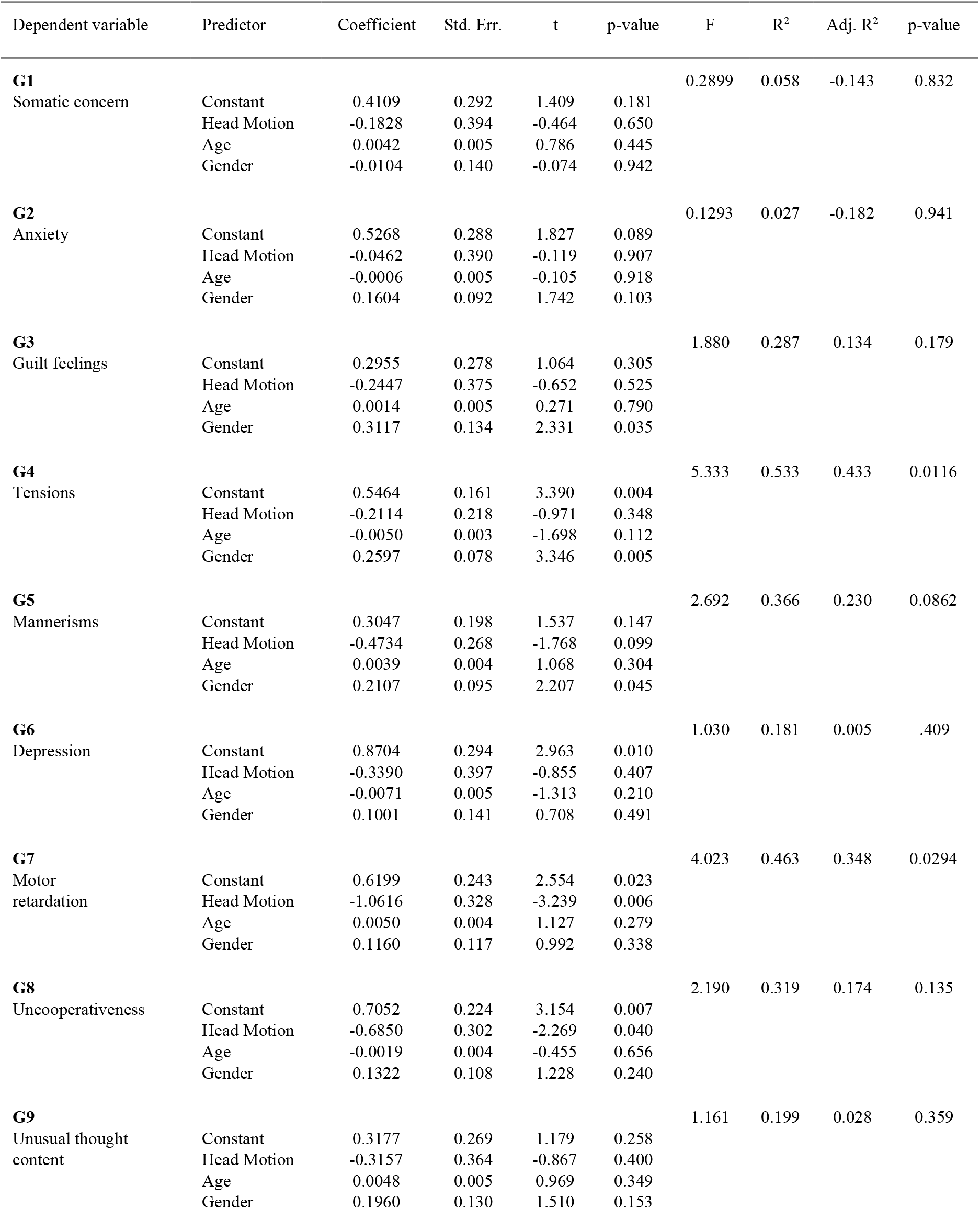
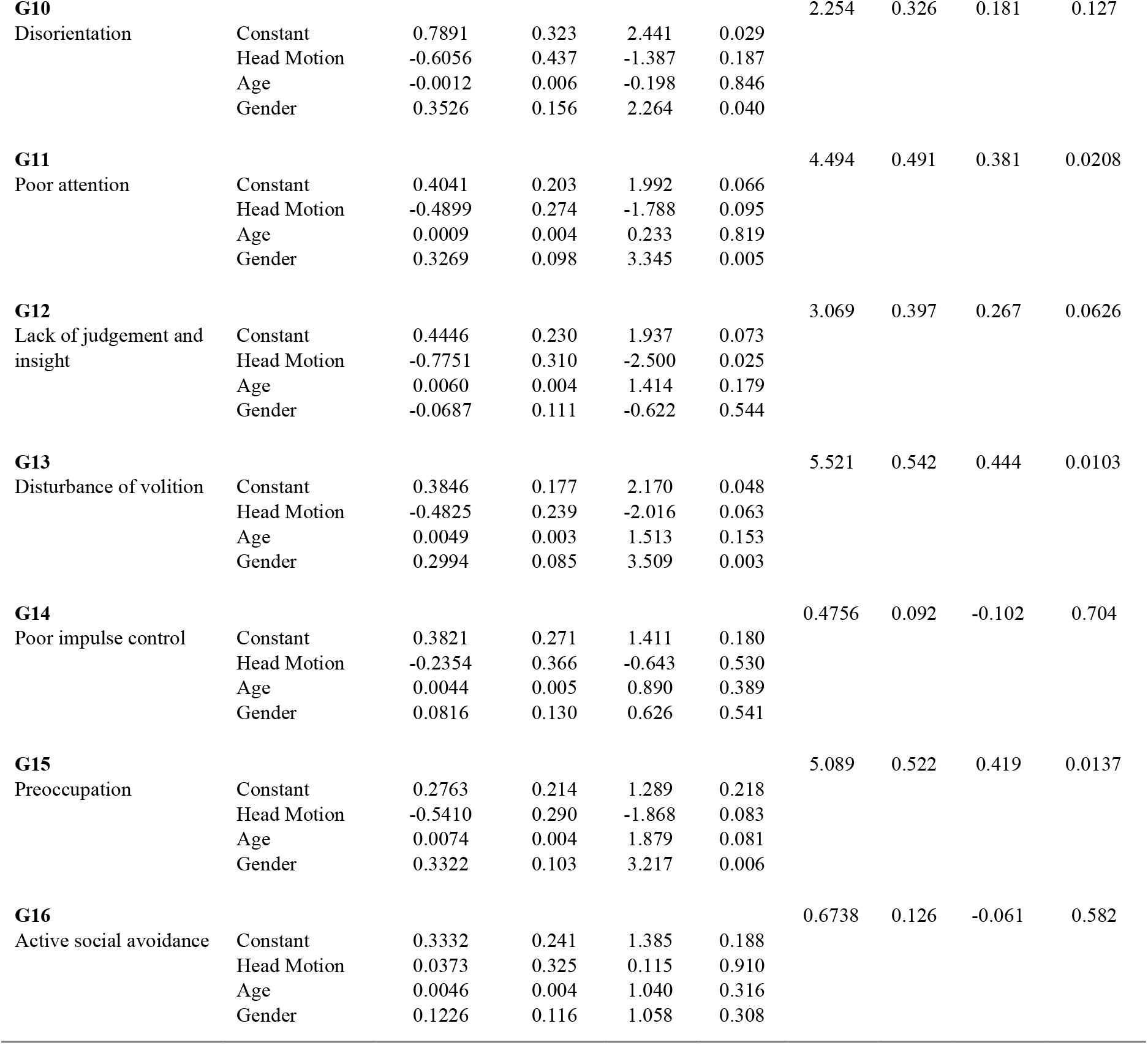
Results for the multiple linear regressions conducted with head movement, age, and gender as predictors of individual items on the PANSS G subscale

| Dependent variable | Predictor | Coefficient | Std. Err. | t | p-value | F | R<sup>2</sup> | Adj. R<sup>2</sup> | p-value |
| --- | --- | --- | --- | --- | --- | --- | --- | --- | --- |
| **G1** |  |  |  |  |  | 0.2899 | 0.058 | -0.143 | 0.832 |
| Somatic concern | Constant | 0.4109 | 0.292 | 1.409 | 0.181 |  |  |  |  |
|  | Head Motion | -0.1828 | 0.394 | -0.464 | 0.650 |  |  |  |  |
|  | Age | 0.0042 | 0.005 | 0.786 | 0.445 |  |  |  |  |
|  | Gender | -0.0104 | 0.140 | -0.074 | 0.942 |  |  |  |  |
| **G2** |  |  |  |  |  | 0.1293 | 0.027 | -0.182 | 0.941 |
| Anxiety | Constant | 0.5268 | 0.288 | 1.827 | 0.089 |  |  |  |  |
|  | Head Motion | -0.0462 | 0.390 | -0.119 | 0.907 |  |  |  |  |
|  | Age | -0.0006 | 0.005 | -0.105 | 0.918 |  |  |  |  |
|  | Gender | 0.1604 | 0.092 | 1.742 | 0.103 |  |  |  |  |
| **G3** |  |  |  |  |  | 1.880 | 0.287 | 0.134 | 0.179 |
| Guilt feelings | Constant | 0.2955 | 0.278 | 1.064 | 0.305 |  |  |  |  |
|  | Head Motion | -0.2447 | 0.375 | -0.652 | 0.525 |  |  |  |  |
|  | Age | 0.0014 | 0.005 | 0.271 | 0.790 |  |  |  |  |
|  | Gender | 0.3117 | 0.134 | 2.331 | 0.035 |  |  |  |  |
| **G4** |  |  |  |  |  | 5.333 | 0.533 | 0.433 | 0.0116 |
| Tensions | Constant | 0.5464 | 0.161 | 3.390 | 0.004 |  |  |  |  |
|  | Head Motion | -0.2114 | 0.218 | -0.971 | 0.348 |  |  |  |  |
|  | Age | -0.0050 | 0.003 | -1.698 | 0.112 |  |  |  |  |
|  | Gender | 0.2597 | 0.078 | 3.346 | 0.005 |  |  |  |  |
| **G5** |  |  |  |  |  | 2.692 | 0.366 | 0.230 | 0.0862 |
| Mannerisms | Constant | 0.3047 | 0.198 | 1.537 | 0.147 |  |  |  |  |
|  | Head Motion | -0.4734 | 0.268 | -1.768 | 0.099 |  |  |  |  |
|  | Age | 0.0039 | 0.004 | 1.068 | 0.304 |  |  |  |  |
|  | Gender | 0.2107 | 0.095 | 2.207 | 0.045 |  |  |  |  |
| **G6** |  |  |  |  |  | 1.030 | 0.181 | 0.005 | .409 |
| Depression | Constant | 0.8704 | 0.294 | 2.963 | 0.010 |  |  |  |  |
|  | Head Motion | -0.3390 | 0.397 | -0.855 | 0.407 |  |  |  |  |
|  | Age | -0.0071 | 0.005 | -1.313 | 0.210 |  |  |  |  |
|  | Gender | 0.1001 | 0.141 | 0.708 | 0.491 |  |  |  |  |
| **G7** |  |  |  |  |  | 4.023 | 0.463 | 0.348 | 0.0294 |
| Motor retardation | Constant | 0.6199 | 0.243 | 2.554 | 0.023 |  |  |  |  |
|  | Head Motion | -1.0616 | 0.328 | -3.239 | 0.006 |  |  |  |  |
|  | Age | 0.0050 | 0.004 | 1.127 | 0.279 |  |  |  |  |
|  | Gender | 0.1160 | 0.117 | 0.992 | 0.338 |  |  |  |  |
| **G8** |  |  |  |  |  | 2.190 | 0.319 | 0.174 | 0.135 |
| Uncooperativeness | Constant | 0.7052 | 0.224 | 3.154 | 0.007 |  |  |  |  |
|  | Head Motion | -0.6850 | 0.302 | -2.269 | 0.040 |  |  |  |  |
|  | Age | -0.0019 | 0.004 | -0.455 | 0.656 |  |  |  |  |
|  | Gender | 0.1322 | 0.108 | 1.228 | 0.240 |  |  |  |  |
| **G9** |  |  |  |  |  | 1.161 | 0.199 | 0.028 | 0.359 |
| Unusual thought content | Constant | 0.3177 | 0.269 | 1.179 | 0.258 |  |  |  |  |
|  | Head Motion | -0.3157 | 0.364 | -0.867 | 0.400 |  |  |  |  |
|  | Age | 0.0048 | 0.005 | 0.969 | 0.349 |  |  |  |  |
|  | Gender | 0.1960 | 0.130 | 1.510 | 0.153 |  |  |  |  |
| **G10** |  |  |  |  |  | 2.254 | 0.326 | 0.181 | 0.127 |
| Disorientation | Constant | 0.7891 | 0.323 | 2.441 | 0.029 |  |  |  |  |
|  | Head Motion | -0.6056 | 0.437 | -1.387 | 0.187 |  |  |  |  |
|  | Age | -0.0012 | 0.006 | -0.198 | 0.846 |  |  |  |  |
|  | Gender | 0.3526 | 0.156 | 2.264 | 0.040 |  |  |  |  |
| **G11** |  |  |  |  |  | 4.494 | 0.491 | 0.381 | 0.0208 |
| Poor attention | Constant | 0.4041 | 0.203 | 1.992 | 0.066 |  |  |  |  |
|  | Head Motion | -0.4899 | 0.274 | -1.788 | 0.095 |  |  |  |  |
|  | Age | 0.0009 | 0.004 | 0.233 | 0.819 |  |  |  |  |
|  | Gender | 0.3269 | 0.098 | 3.345 | 0.005 |  |  |  |  |
| **G12** |  |  |  |  |  | 3.069 | 0.397 | 0.267 | 0.0626 |
| Lack of judgement and insight | Constant | 0.4446 | 0.230 | 1.937 | 0.073 |  |  |  |  |
|  | Head Motion | -0.7751 | 0.310 | -2.500 | 0.025 |  |  |  |  |
|  | Age | 0.0060 | 0.004 | 1.414 | 0.179 |  |  |  |  |
|  | Gender | -0.0687 | 0.111 | -0.622 | 0.544 |  |  |  |  |
| **G13** |  |  |  |  |  | 5.521 | 0.542 | 0.444 | 0.0103 |
| Disturbance of volition | Constant | 0.3846 | 0.177 | 2.170 | 0.048 |  |  |  |  |
|  | Head Motion | -0.4825 | 0.239 | -2.016 | 0.063 |  |  |  |  |
|  | Age | 0.0049 | 0.003 | 1.513 | 0.153 |  |  |  |  |
|  | Gender | 0.2994 | 0.085 | 3.509 | 0.003 |  |  |  |  |
| **G14** |  |  |  |  |  | 0.4756 | 0.092 | -0.102 | 0.704 |
| Poor impulse control | Constant | 0.3821 | 0.271 | 1.411 | 0.180 |  |  |  |  |
|  | Head Motion | -0.2354 | 0.366 | -0.643 | 0.530 |  |  |  |  |
|  | Age | 0.0044 | 0.005 | 0.890 | 0.389 |  |  |  |  |
|  | Gender | 0.0816 | 0.130 | 0.626 | 0.541 |  |  |  |  |
| **G15** |  |  |  |  |  | 5.089 | 0.522 | 0.419 | 0.0137 |
| Preoccupation | Constant | 0.2763 | 0.214 | 1.289 | 0.218 |  |  |  |  |
|  | Head Motion | -0.5410 | 0.290 | -1.868 | 0.083 |  |  |  |  |
|  | Age | 0.0074 | 0.004 | 1.879 | 0.081 |  |  |  |  |
|  | Gender | 0.3322 | 0.103 | 3.217 | 0.006 |  |  |  |  |
| **G16** |  |  |  |  |  | 0.6738 | 0.126 | -0.061 | 0.582 |
| Active social avoidance | Constant | 0.3332 | 0.241 | 1.385 | 0.188 |  |  |  |  |
|  | Head Motion | 0.0373 | 0.325 | 0.115 | 0.910 |  |  |  |  |
|  | Age | 0.0046 | 0.004 | 1.040 | 0.316 |  |  |  |  |
|  | Gender | 0.1226 | 0.116 | 1.058 | 0.308 |  |  |  |  |

